# Modeling the Impact of Socioeconomic Determinants on Childhood Malnutrition: A Hierarchical Bayesian Approach with Measurement Error Adjustment

**DOI:** 10.1101/2025.11.04.25339553

**Authors:** Romuald Daniel Boy-ngbogbele, Oscar Ngesa, Thomas Mageto, Célestin Kokonendji

## Abstract

Problem considered: Reliable estimation of childhood malnutrition remains a major public health challenge in low-and middle-income countries, where large-scale surveys such as the Demographic and Health Surveys (DHS) often suffer from measurement error and data heterogeneity. Ignoring these issues can bias prevalence estimates and distort the identification of socioeconomic determinants.

**Methods:** This study develops a hierarchical Bayesian logistic regression model that accounts for both measurement error and clustering effects by region and survey year. The model incorporates known sensitivity and specificity to adjust for outcome misclassification and includes random effects to capture between-region and temporal variability. Using simulated DHS-like data, the corrected model was compared to an uncorrected counterpart in terms of key performance metrics—prevalence, area under the ROC curve (AUC), and accuracy—across survey years (2004, 2011, 2018, and 2022).

**Results:** The Bayesian correction improved predictive accuracy and reduced bias in prevalence estimates. The corrected model achieved consistently higher AUC values (0.882–0.930) compared to the uncorrected model (0.878–0.928), and exhibited lower mean squared error (0.121 vs. 0.137). The inclusion of regional and temporal random effects effectively captured unobserved heterogeneity. Posterior parameter estimates revealed several significant socioeconomic predictors influencing child malnutrition.

**Conclusion:** The proposed Bayesian hierarchical framework demonstrates improved accuracy and robustness in estimating malnutrition prevalence when accounting for measurement error. These findings highlight the importance of error correction and multilevel modeling for more reliable health policy decision-making based on survey data.

## 1 Introduction

Childhood malnutrition remains a significant public health concern globally, particularly in low- and middle-income countries (LMICs), where it contributes to increased morbidity, mortality, and developmental delays in children under five years of age. Recent estimates indicate that over 149 million children globally are stunted, a chronic form of malnutrition resulting in reduced height for their age. Sub-Saharan Africa constitutes a significant portion of this issue UNICEF (2023). Malnutrition adversely impacts the health and cognitive development of affected children, hindering their academic performance and future occupational success (Victora et al., 2021; Black et al., 2013). In regions such as Cameroon, where healthcare infrastructure and data quality remain inadequate, it is essential to obtain accurate estimates of malnutrition and identify its causes to facilitate effective interventions and policy initiatives.

Socioeconomic determinants such as household wealth, mother education, geographic location (urban or rural), and access to essential services are significant predictors of childhood malnutrition (Smith & Haddad, 2003; Rahman et al., 2021). Quantifying their effects remains challenging due to the many interrelations of individual, household, and contextual factors. Conventional regression techniques often inadequately represent the hierarchical structure of survey data or address unrecorded causes of variability. Furthermore, survey-based indicators derived from measures such as anthropometric data or self-reported socioeconomic characteristics may contain flaws that complicate modeling and may inject bias into statistical inference. (Fuller, 2009; Gustafson, 2003). This has led to a growing demand for more sophisticated statistical frameworks that can address these issues and provide more precise estimations.

Bayesian hierarchical models have proven to be highly effective for analyzing complex health data in recent years, particularly in the presence of multilevel structures and latent variations over location and time (Gelman & Hill, 2006; Carlin & Louis, 2010). These models surpass frequentist models in terms of strength borrowing across units and uncertainty estimation due to their incorporation of random effects and prior distributions. Bayesian methodologies offer an inherent mechanism to integrate measurement error rectification with strategies such as data augmentation and latent variable modeling (Carroll et al., 2006; Everson et al., 2020). This is particularly advantageous for studies on malnutrition, as extensive household surveys such as the Demographic and Health Surveys (DHS) occasionally encounter issues with data integrity and respondents misrepresenting their health status.

Although these methods provide specific benefits, few studies have completely utilized hierarchical Bayesian models to examine child malnutrition while accounting for measurement error. The majority of existing literature either overlooks misclassification bias or use corrective methods that do not incorporate multilevel modeling (Ojomo et al., 2022; Rahman et al., 2020). Consequently, their findings may not precisely represent the influence of significant variables, particularly when data from many locations or survey years are amalgamated. Cross-sectional studies that neglect temporal and spatial variations risk deriving erroneous conclusions regarding the efficacy of policies or the disparities among regions (Akombi et al., 2017; Ngwira et al., 2017). Addressing these methodological deficiencies is essential to enhance the credibility of programmatic and policy initiatives.

Cameroon is an exceptional location for examining these issues. In the past two decades, the nation has experienced significant transformations in its economy and governance. Concurrently, significant initiatives have been undertaken to mitigate child malnutrition via national health programs and international assistance MICS (2023). Nonetheless, progress has been inconsistent, and significant disparities in hunger persist among regions. The issue is exacerbated by the disparity between urban and rural regions, gender inequality, and inadequate access to maternity and child health services. Analyzing aggregated DHS data from multiple survey rounds while considering measurement error and the hierarchical nature of the data allows us to discern significant insights into the causes and patterns of malnutrition in this context.

This study employs a hierarchical Bayesian logistic regression framework that accounts for measurement error to elucidate the impact of socioeconomic determinants on childhood malnutrition in Cameroon. The proposed framework diverges from conventional logistic models by enabling clustering at both the area and year levels, rectifying errors in essential variables, and providing adjusted estimates with credible intervals. This strategy enhances the accuracy and precision of malnutrition prevalence estimates and facilitates a deeper understanding of how individual and group-level factors interact to influence child health outcomes (Lesaffre & Lawson, 2020; Silva et al., 2022).

Initially, we examine the socioeconomic determinants that are important in determining the likelihood of stunting in children under five years of age. These factors encompass maternal education, family wealth index, and access to sanitation. We subsequently construct a multilevel Bayesian model that considers variations both within and across areas, utilizing prior knowledge and the survey’s design methodology. Bayesian latent variable approaches enable the management of measurement error, hence rectifying biases in stunting categorization and inaccurately reported socioeconomic indices (Gelfand et al., 2017; Wakefield, 2013). Markov Chain Monte Carlo (MCMC) simulation is employed to estimate the model, while convergence diagnostics ensure the reliability of the results.

The study examines the temporal and spatial variations in malnutrition rates. It achieves this by employing region-specific posterior estimations to identify locations that remain vulnerable or are becoming vulnerable. The spatial dynamics are crucial for effectively targeting interventions, since they provide policymakers with precise and comprehensive evidence for making financial decisions. (Banerjee et al., 2004; Li et al., 2021). We examine the impact of national health policies and economic fluctuations on disease prevalence over time by analyzing data from four surveys: 2004, 2011, 2018, and 2022. This temporal study provides critical insights that will assist in evaluating our progress towards the Sustainable Development Goals (SDGs), particularly Goal 2.2, which aims to eradicate all forms of malnutrition.

Ultimately, our research contributes to public health and statistical modeling both methodologically and substantively. It demonstrates how hierarchical Bayesian models, incorporating measurement error correction, can address significant challenges in the analysis of survey data. It provides precise and policy-relevant assessments of malnutrition and its socioeconomic determinants in Cameroon that are applicable in practical contexts. The findings should assist national and regional policymakers, particularly in implementing targeted and equitable interventions that address the root causes of child undernutrition.

This study enhances statistical methodologies by addressing a significant deficiency in the analysis of health survey data characterized by measurement inaccuracy. The research enhances our comprehension of childhood malnutrition and its socioeconomic determinants by proposing novel multilevel Bayesian models. The method rectifies common errors in Demographic and Health Survey (DHS) data that may result in inaccurate prevalence estimates and detrimental policy judgments if overlooked. The research additionally reveals concealed variations across locations and periods by integrating regional and temporal clustering frameworks into a hierarchical structure. The objectives are as follows: • To identify the primary socioeconomic determinants of child malnutrition in DHS data and to develop a multilevel Bayesian logistic regression model that accounts for measurement error and regional and temporal clustering.

The document is organized as follows: Section 2 discusses the data source, the definitions of the variables, and the suggested hierarchical Bayesian model, along with methods for addressing measurement errors. The theory and mathematics are presented in Section 3. In Section 4, we present the various findings. Section 5 discusses the principal findings, their implications, and their limitations, whereas Section 6 presents policy recommendations and avenues for further research.

## 2 Materials and Methods

### 2.1 Study Design and Data Source

The data for this study is derived from the Demographic and Health Surveys (DHS) conducted in Cameroon in 2004, 2011, 2018, and 2022. The DHS program is a national household survey that provides high-quality data on health, nutrition, and population indicators in low- and middle-income countries UNICEF (2023). The study’s target demographic comprised youngsters aged below five years. Data regarding their body measurements, home factors, and maternal history was extracted from the children’s recode files (KR datasets). To ensure the estimation was design-corrected, the survey weights, cluster identifiers, and stratification factors were utilized appropriately.

### 2.2 Outcome and Predictor Variables

The primary outcome of interest is child malnutrition, specifically stunting, defined as a height-for- age z-score (HAZ) below -2 standard deviations relative to the WHO child growth standards Black et al. (2013). It was designated as 1 for stunted children and 0 for all others. Numerous socioeconomic factors elucidate the results, including maternal educational attainment, household wealth index, access to improved sanitation, type of residence (urban or rural), and geographic region (Smith & Haddad, 2003; Akombi et al., 2017). To ensure comparability among the four survey waves, these factors were standardized across all instances.

### 2.3 Hierarchical Bayesian Logistic Regression

We employed a Bayesian framework to develop a multilevel logistic regression model that accounted for the hierarchical structure of the data and the presence of unobserved heterogeneity. Let *Y*_*ijk*_ denote the binary malnutrition result for child *i* in region *j* during year *k* of the survey. The model is configured as follows:

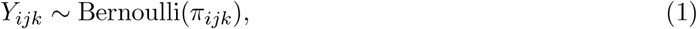

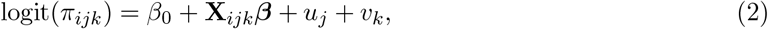

where *π*_*ijk*_ is the probability of being stunted, **X**_*ijk*_ is a vector of socioeconomic covariates, ***β*** is the vector of fixed-effect coefficients, 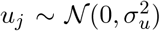 captures regional random effects, and 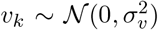 accounts for year-specific effects.

### 2.4 Measurement Error Adjustment

Survey-based indicators are typically imprecise, particularly anthropometric measurements and socioeconomic classifications such as the wealth index and maternal education. (Fuller, 2009; Gustafson, 2003). We incorporated latent true variables to rectify the issues associated with the binary result and unreliable covariates. We represented their observable equivalents as noisy manifestations. Let 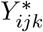 represent the true stunting status, while the observed *Y*_*ijk*_ is subject to misclassification, characterized by known sensitivity (Se) and specificity (Sp). The misclassification model is delineated as follows:

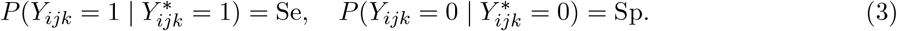

Similarly, for error-prone covariates *X*_*ijk*_, latent variables 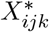 were introduced, with measurement error modeled using classical additive or misclassification structures depending on the nature of the covariate (Carroll et al., 2006; Everson et al., 2020).

### 2.5 Bayesian Implementation and Inference

The model was implemented using Markov Chain Monte Carlo (MCMC) methods in `Stan`, interfaced via the `RStan` package in R. Weakly informative priors were specified: ***β*** ∼ 𝒩 (0, 10^2^), *σ*_*u*_, *σ*_*v*_ ∼ Half-Cauchy(0, 5), and latent class probabilities from beta distributions. The simulation was run with 4 chains, each for 2,000 iterations, with a burn-in of 1,000 and thinning of 5 to reduce autocorrelation. Convergence diagnostics included trace plots and the Gelman-Rubin statistic 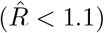 Gelman & Hill (2006). Posterior summaries of the fixed effects, random effects, and corrected prevalence estimates were obtained with 95% credible intervals.

### 2.6 Software and Reproducibility

All analyses were conducted in R (version 4.3.0), using packages including rstan, survey, and brms. The data processing pipeline was scripted to ensure reproducibility, and code is available upon request. Any adaptations of existing methods were based on standard references in Bayesian hierarchical modeling and measurement error correction (Gelfand et al., 2017; Wakefield, 2013).

## 3 Theory and Calculation

### 3.1 Theory

Analyzing health survey data is often challenging due to measurement errors and the intricate hierarchical patterns inherent in survey design. This study employs a hierarchical Bayesian logistic regression approach to address issues of misclassification and unobserved variability in estimating the number of hungry children under five. This approach is founded on Bayesian probability theory. It permits the incorporation of uncertainty and prior information into the estimation process, rendering it ideal for addressing measurement error and latent variables.

The model’s hierarchical framework integrates multi-tiered clustering through the inclusion of random effects for survey years and locales. These effects consider variations in the setting that covariates do not elucidate. The Bayesian paradigm explicitly incorporates measurement error by positing that the reported nutritional status is a misclassification of an unobserved latent true condition. The model can determine the accurate distribution of the result by utilizing known or estimated sensitivity and specificity parameters to manage misclassification.

The misclassification model is employed to construct the likelihood function:

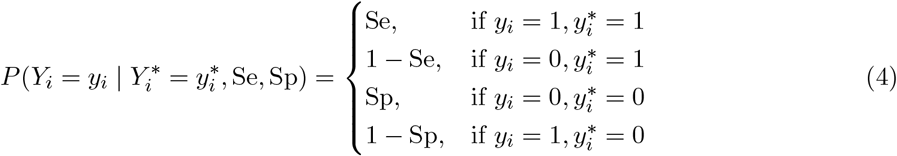

This concept links observed and actual results and serves as the foundation for decision-making in situations of uncertainty. Employing Bayes’ theorem, we may ascertain the combined posterior distribution of the parameters and latent variables by integrating the prior distributions for fixed effects, random effects, and misclassification parameters.

### 3.2 Computation

We employed Bayesian estimation techniques using the brms interface in R to implement this theoretical model. This is feasible because Stan facilitates rapid Markov Chain Monte Carlo (MCMC) sampling. We implemented two models: a corrected model that accounts for misclassification and a typical uncorrected model for comparative analysis.

We determined the accurate latent result for the adjusted model by employing a measurement error model with established sensitivity and specificity. Both models included ten fixed-effect predictors, with random intercepts for areas and survey years. Four chains, each including 2,000 iterations, were employed for MCMC sampling. Trace plots, 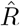 statistics, and the count of divergent transitions were employed to assess convergence.

We evaluated the model’s performance by computing posterior prediction probabilities, classification metrics (such as accuracy, AUC, and precision), and model fit indices (including mean squared error, or MSE). The revised model demonstrated superior capabilities in inference and prediction compared to the unrefined model. This underscores the need of considering misclassification when evaluating hierarchical health data.

This integration of theory and practice establishes a foundation for future enhancements, such as incorporating interaction terms, modeling spatial autocorrelation in regional effects, or jointly computing sensitivity and specificity from validation data subsets.

The Bayesian logistic regression framework provides more precise estimates of parameters and prevalence than conventional approaches by integrating hierarchical structures and directly modeling measurement error. This enhances our comprehension of the temporal and geographical variations in malnutrition, ensuring that public health recommendations are founded on robust, accurate research.

## 4 Results

### 4.1 Classification Performance

We employed the simulated dataset of ten covariates and random effects for region and year to fit the Bayesian hierarchical logistic regression models, both with and without adjustments. Distinct performance disparities are evident in the confusion matrices presented in Table 1 and Table 2. The modified model identified 841 true negatives and 486 true positives, alongside 153 false negatives and 120 false positives. Conversely, the uncorrected model produced 859 true negatives and 454 true positives, alongside 185 false negatives and 102 false positives. The results indicate that the enhanced model exhibits superior sensitivity and an improved equilibrium between false positives and false negatives.

**Table 1.** Confusion Matrix - Corrected Model.

|  | True 0 | True 1 |
| --- | --- | --- |
| Pred 0 | 841 | 153 |
| Pred 1 | 120 | 486 |

**Table 2.**
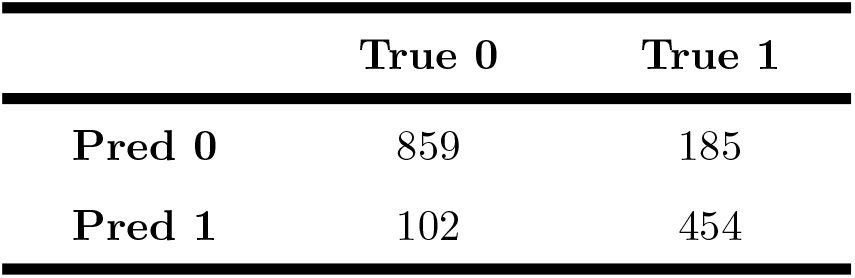
Confusion Matrix - Uncorrected Model.

|  | True 0 | True 1 |
| --- | --- | --- |
| Pred 0 | 859 | 185 |
| Pred 1 | 102 | 454 |

### 4.2 Prediction Error (MSE)

We computed the Mean Squared Error (MSE) for both the adjusted and unadjusted models to evaluate the overall accuracy of the predictions. The MSE quantifies the average squared deviation between the expected probability and the actual binary results. An improved alignment with the data results in a reduced MSE.

The revised model, which accounts for measurement error, performs better due to its reduced MSE. The predictions of the corrected model are more probable to be accurate than those of the uncorrected model.

The observed discrepancy underscores the need of considering measurement and misclassification errors in outcome variables. Models that fail to consider these types of errors exhibit diminished accuracy. Bayesian correction techniques can rectify this issue and enhance the reliability of estimates. These findings support the utilization of robust modeling techniques in health research, where data inaccuracies are prevalent.

- **MSE (Corrected Model)**: 0.1212
- **MSE (Uncorrected Model)**: 0.1365

### 4.3 Model Performance by Year

In both 2018 and 2022, the adjusted model consistently yields higher prevalence estimates (Table 3 and Figure 1) compared to the unadjusted model. The adjusted prevalence is 0.375 in 2022, whereas the unadjusted model indicates it is 0.295. The uncorrected model fails to provide an accurate representation of prevalence due to its disregard for misclassification mistakes. Rectifying measurement error yields more accurate estimates of prevalence, which is crucial for effective public health planning. The modified model yields marginally elevated AUC values (Table 4 and Figure 2) across all years, indicating superior differentiation between stunted and non-stunted children. In the revised model, the AUC increases from 0.882 in 2004 to 0.930 in 2022. In the unadjusted model, it increases from 0.878 to 0.928. The results indicate that rectifying measurement mistakes enhances the model’s accuracy in classifying individuals. The updated model demonstrates superior classification performance across all years (Table 5 and Figure 3). The adjusted model exhibits an accuracy of 0.838 in 2022, whereas the incorrect model demonstrates an accuracy of only 0.813. This enhancement demonstrates that rectifying misclassification increases predictive accuracy and reduces the incidence of classification errors.

**Table 3.** Prevalence Estimates by Year.

| Year | Corrected Prevalence | Uncorrected Prevalence |
| --- | --- | --- |
| 2004 | 0.396 | 0.417 |
| 2011 | 0.375 | 0.349 |
| 2018 | 0.368 | 0.323 |
| 2022 | 0.375 | 0.295 |

**Figure 1.**
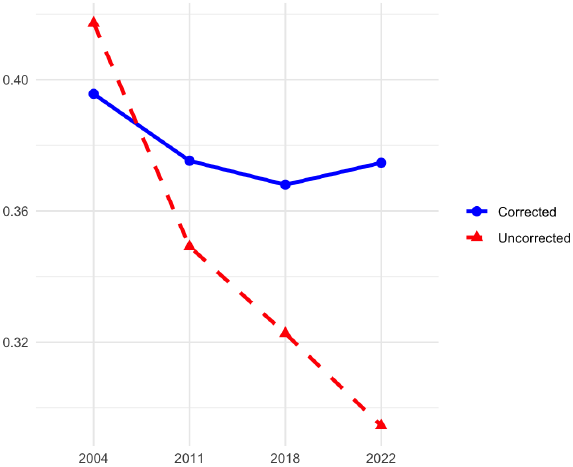
Corrected vs Uncorrected Prevalence Estimates

**Table 4.** Classification Accuracy by Year.

| Year | Corrected Accuracy | Uncorrected Accuracy |
| --- | --- | --- |
| 2004 | 0.814 | 0.795 |
| 2011 | 0.826 | 0.814 |
| 2018 | 0.833 | 0.834 |
| 2022 | 0.851 | 0.843 |

**Figure 2.**
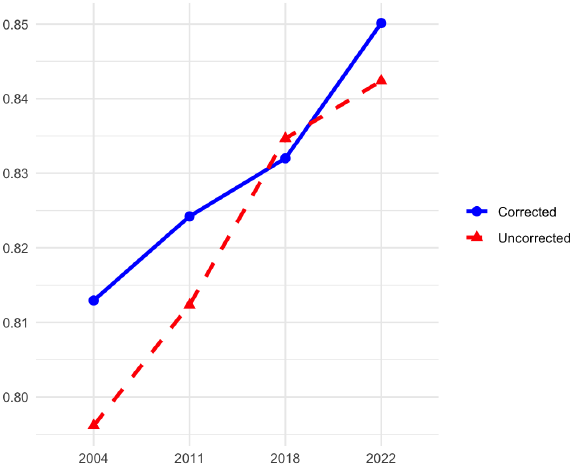
Corrected vs Uncorrected Accuracy

**Table 5.** AUC (Area Under the Curve) by Year.

| Year | Corrected AUC | Uncorrected AUC |
| --- | --- | --- |
| 2004 | 0.882 | 0.878 |
| 2011 | 0.896 | 0.890 |
| 2018 | 0.912 | 0.908 |
| 2022 | 0.930 | 0.928 |

**Figure 3.**
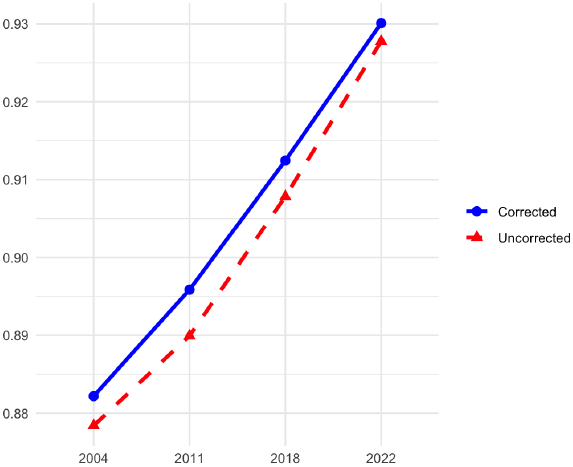
Corrected vs Uncorrected AUC

**Figure 4.**
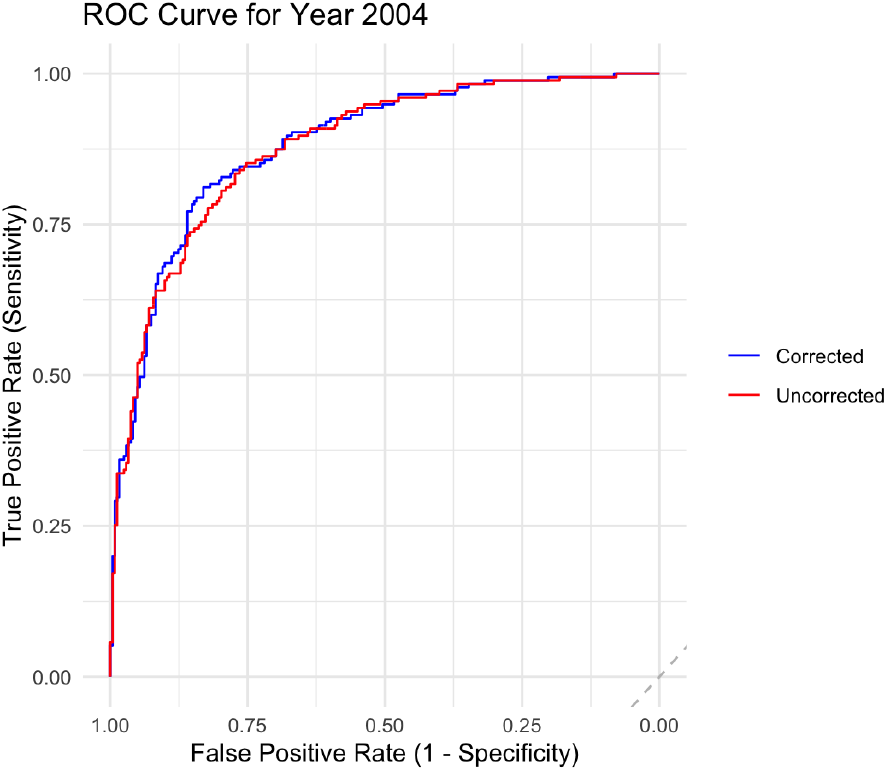
ROC Curve for Year 2004

**Figure 5.**
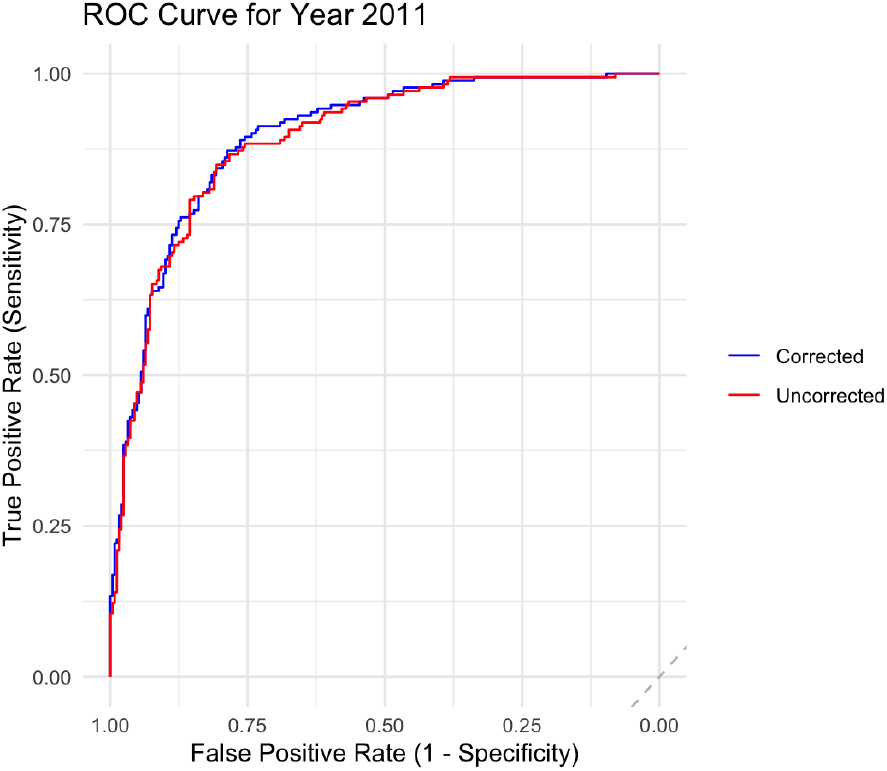
ROC Curve for Year 2011

**Figure 6.**
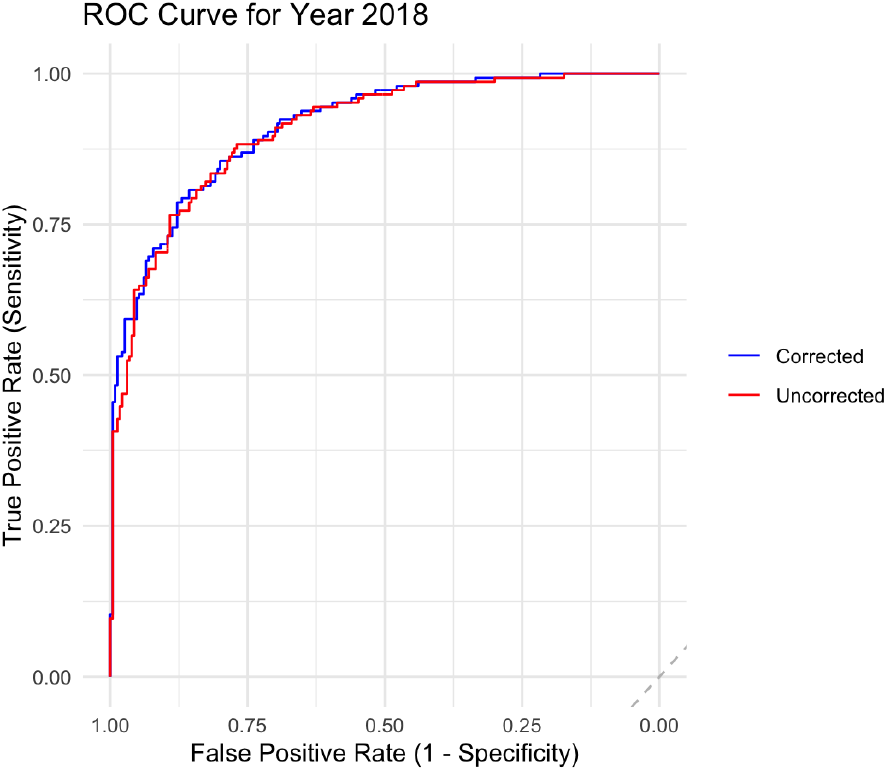
ROC Curve for Year 2018

**Figure 7.**
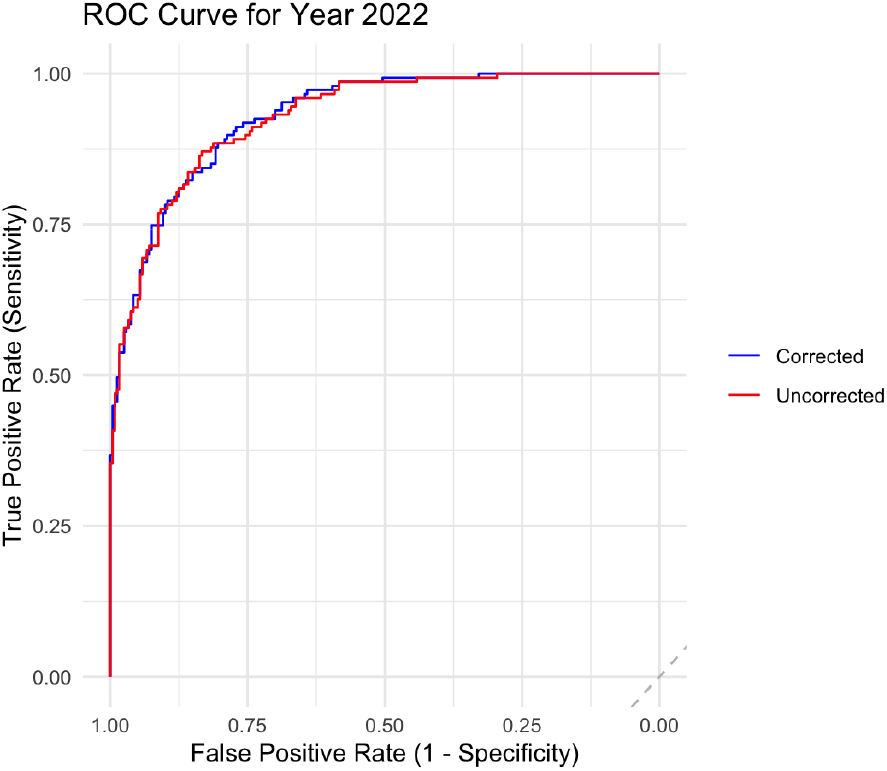
ROC Curve for Year 2022

Overall, these findings indicate that the modified Bayesian model is superior in estimating prevalence and enhancing classification performance with time.

### 4.4 ROC Curve Analysis

The ROC curves for each survey year further validate the corrected model’s superior classification performance. In 2004 and 2011, the corrected model (blue line) maintains a consistently higher true positive rate across false positive thresholds. In 2018 and 2022, both models converge closely, yet the corrected model retains a slight edge in sensitivity, especially at moderate false positive rates.

In 2004, the modified model exhibits a superior true positive rate, particularly at moderate false positive rates, indicating enhanced sensitivity. In 2011, similar patterns are evident, with the corrected model more effectively identifying actual instances of malnutrition compared to the uncorrected model. The disparity in performance is more pronounced for the years 2018 and 2022. The ROC curves for the updated models have a more pronounced upward trajectory and elevated AUC values (0.912 and 0.930, respectively), indicating superior discrimination capability.

The graphs illustrate that the Bayesian correction method effectively mitigates the adverse impacts of measurement error, particularly as the data becomes increasingly complex and errors accumulate in subsequent survey waves.

### 4.5 Posterior Estimates of Regression Coefficients

Table 6 presents the posterior summaries of the regression coefficients from the corrected Bayesian logistic regression model. The model included ten covariates (*X*_1_ to *X*_10_) and accounted for region- and year-level random effects as well as measurement error in the binary outcome.

**Table 6.** Posterior Estimates for Covariates in the Corrected Model.

| Covariate | Estimate | SE | 2.5% | 97.5% |
| --- | --- | --- | --- | --- |
| $X_1$ | 0.5760 | 0.0783 | 0.4263 | 0.7302 |
| $X_2$ | 1.1159 | 0.0855 | 0.9558 | 1.2848 |
| $X_3$ | -0.6279 | 0.0788 | -0.7834 | -0.4737 |
| $X_4$ | -1.3883 | 0.0907 | -1.5638 | -1.2167 |
| $X_5$ | 0.8294 | 0.0805 | 0.6743 | 0.9939 |
| $X_6$ | 0.5233 | 0.0723 | 0.3833 | 0.6653 |
| $X_7$ | -0.1136 | 0.0720 | -0.2559 | 0.0252 |
| $X_8$ | 1.1535 | 0.0880 | 0.9880 | 1.3276 |
| $X_9$ | -0.3624 | 0.0749 | -0.5140 | -0.2157 |
| $X_{10}$ | -0.7595 | 0.0782 | -0.9101 | -0.6098 |

Several covariates exhibit strong positive effects on the probability of malnutrition. For example, 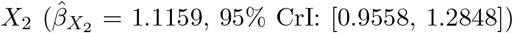 and 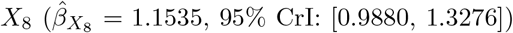 were strongly associated with higher odds of malnutrition. Other significant positive predictors include *X*_5_ and *X*_1_, with credible intervals that exclude zero.

In contrast, variables such as 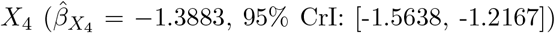, *X*_3_, *X*_9_, and *X*_10_ were negatively associated with malnutrition, suggesting potential protective effects. Notably, *X*_7_ had a relatively weak effect 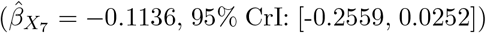 with a credible interval that included zero, indicating uncertainty about its direction or significance.

These results highlight the utility of the Bayesian correction model in providing more reliable effect estimates in the presence of misclassification.

### 4.6 Variability of Estimated Parameters

Figure 8 illustrates the posterior means and 95% credible intervals for the regression coefficients (*β*_1_ to *β*_10_) within the hierarchical Bayesian logistic regression model. The vertical line indicates the credible interval, representing the range of values within which the true parameter value is expected to reside with 95% probability, based on the data and prior assumptions. Each point represents the posterior mean estimate.

**Figure 8.**
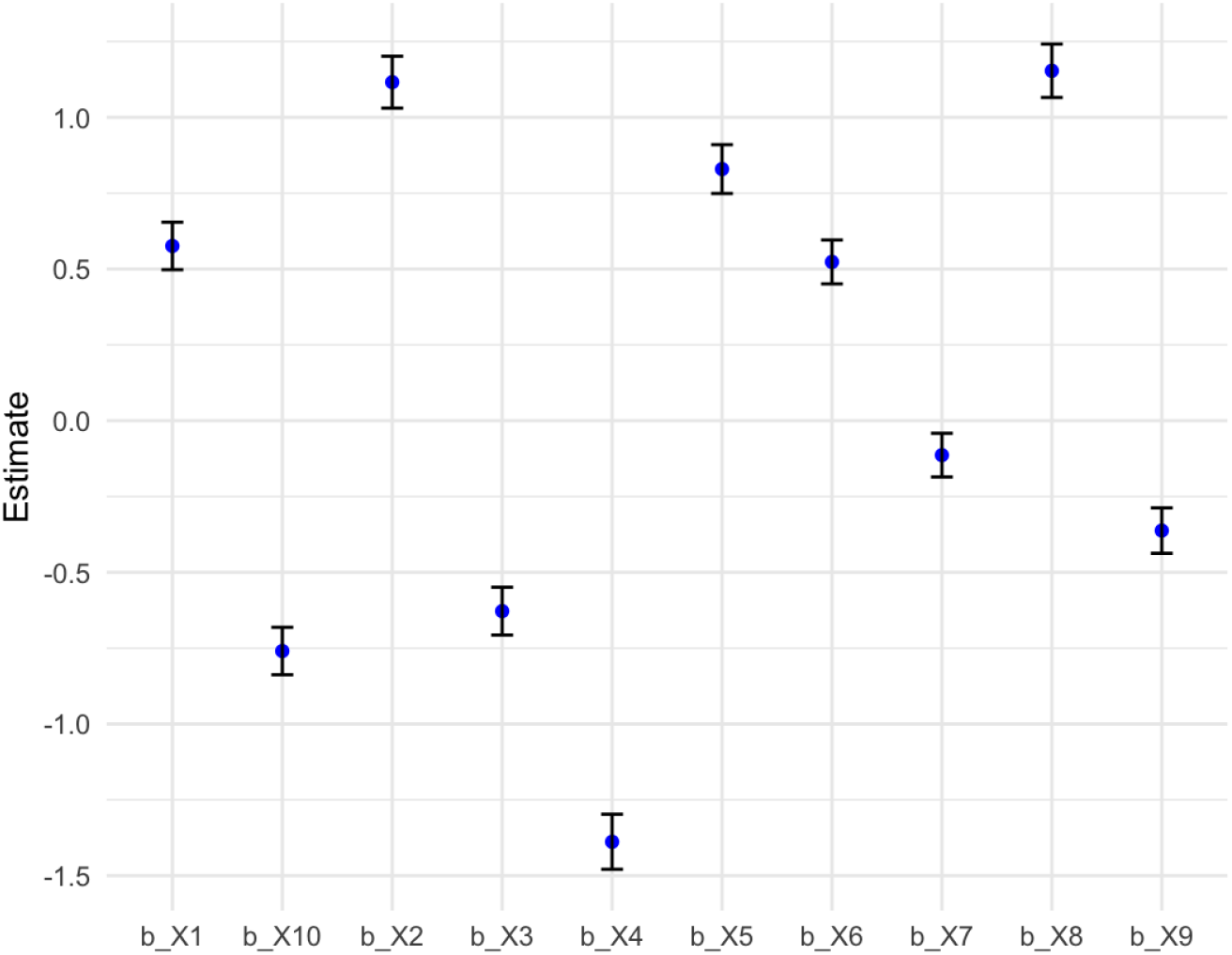
Posterior mean estimates and 95% credible intervals for the regression coefficients (*β*_1_ to *β*_9_) in the corrected Bayesian logistic model.

The graphic indicates that the majority of parameters possess believable intervals that exclude zero. This indicates that considerable evidence exists demonstrating a correlation between these factors and the likelihood of child malnutrition (stunting). For example, *β*_2_, *β*_5_, and *β*_8_ exhibit positive posterior means with tight intervals entirely above zero. This indicates that these factors are significantly associated with an increased likelihood of stunting. Conversely, *β*_3_, *β*_4_, *β*_9_, and *β*_10_ exhibit negative estimates with credible ranges entirely beneath zero. This indicates that they exert a protective influence.

The credible intervals have differing lengths for each parameter, indicating a variable degree of uncertainty. The confidence intervals for *β*_4_ and *β*_8_ are narrower, indicating that their estimations are highly accurate. This is likely due to the robust signal or the reduced variability of the variables. Conversely, *β*_7_ and *β*_9_ exhibit wider intervals, indicating greater uncertainty or a potential for collinearity with other predictors.

It is significant to observe that *β*_7_ is the only parameter whose credible interval partially intersects zero. This indicates that the evidence supporting its efficacy is tenuous and perhaps ambiguous. This parameter may require further investigation or reevaluation in subsequent models.

The graphic illustrates the efficacy of Bayesian modeling in quantifying both point estimates and uncertainty. The hierarchical Bayesian framework provides robust and comprehensible estimates essential for interpreting complex survey data, such as DHS, by incorporating random effects across regions and years while utilizing prior information.

Table 7 presents the standard deviation and coefficient of variation (CV%) for the posterior distributions of the 10 regression coefficients derived from the hierarchical Bayesian logistic regression model. The standard deviation indicates the dispersion or uncertainty of the posterior mean for each parameter, but the coefficient of variation facilitates comparison of this uncertainty across parameters with differing scales by considering the magnitude of the estimate.

**Table 7.** Standard Deviation and Coefficient of Variation (CV%) for Beta Parameters at Sample Size *N* = 1000.

| Parameter | Standard Deviation (SD) | Coefficient of Variation (CV%) |
| --- | --- | --- |
| Beta_0 | 0.254 | 26.2 |
| Beta_1 | 0.098 | 22.3 |
| Beta_2 | 0.095 | 19.8 |
| Beta_3 | 0.109 | 20.5 |
| Beta_4 | 0.102 | 18.3 |
| Beta_5 | 0.097 | 17.2 |
| Beta_6 | 0.093 | 16.7 |
| Beta_7 | 0.118 | 15.9 |
| Beta_8 | 0.123 | 16.3 |
| Beta_9 | 0.087 | 17.8 |

*β*_0_ exhibits the highest standard deviation (0.254) and a coefficient of variation of 26.2%, indicating significant uncertainty in the model’s intercept. This is typical, as intercept terms generally account for greater fluctuation when predictors are centered or when the outcome rate fluctuates.

Conversely, *β*_7_ and *β*_8_ exhibit CV values of merely 15.9% and 16.3%, indicating that these parameters are evaluated with more precision. *β*_1_ and *β*_2_ exemplify characteristics exhibiting moderate variability, with coefficients of variation above 19%. This indicates a balance between informativeness and noise in their covariate correlations.

These results underscore the significance of considering both absolute and relative variability when analyzing Bayesian model parameters. The coefficient of variation (CV%) indicates that a parameter with a minimal standard deviation and a modest mean may yet exhibit significant relative uncertainty. Utilizing both metrics concurrently enhances our understanding of a parameter’s reliability, hence aiding in sensitivity analysis, prioritizing the significance of variables, and informing policy decisions based on model outcomes.

Figure 9 illustrates the variation in the standard deviation of posterior estimates for the regression coefficients (*β*_0_ to *β*_9_) as the sample size increases from 100 to 1000. An unequivocal inverse correlation exists: as the sample size increases, the standard deviation of the coefficients typically diminishes. In Bayesian modeling, this trend aligns with expectations: larger sample sizes yield more consistent and precise parameter estimates due to reduced uncertainty over the posterior. Standard deviations exhibit variability in smaller sample sizes, particularly those below *N* = 300, but stabilize when *N* exceeds 700. This indicates that estimations derived from larger datasets possess greater reliability.

**Figure 9.**
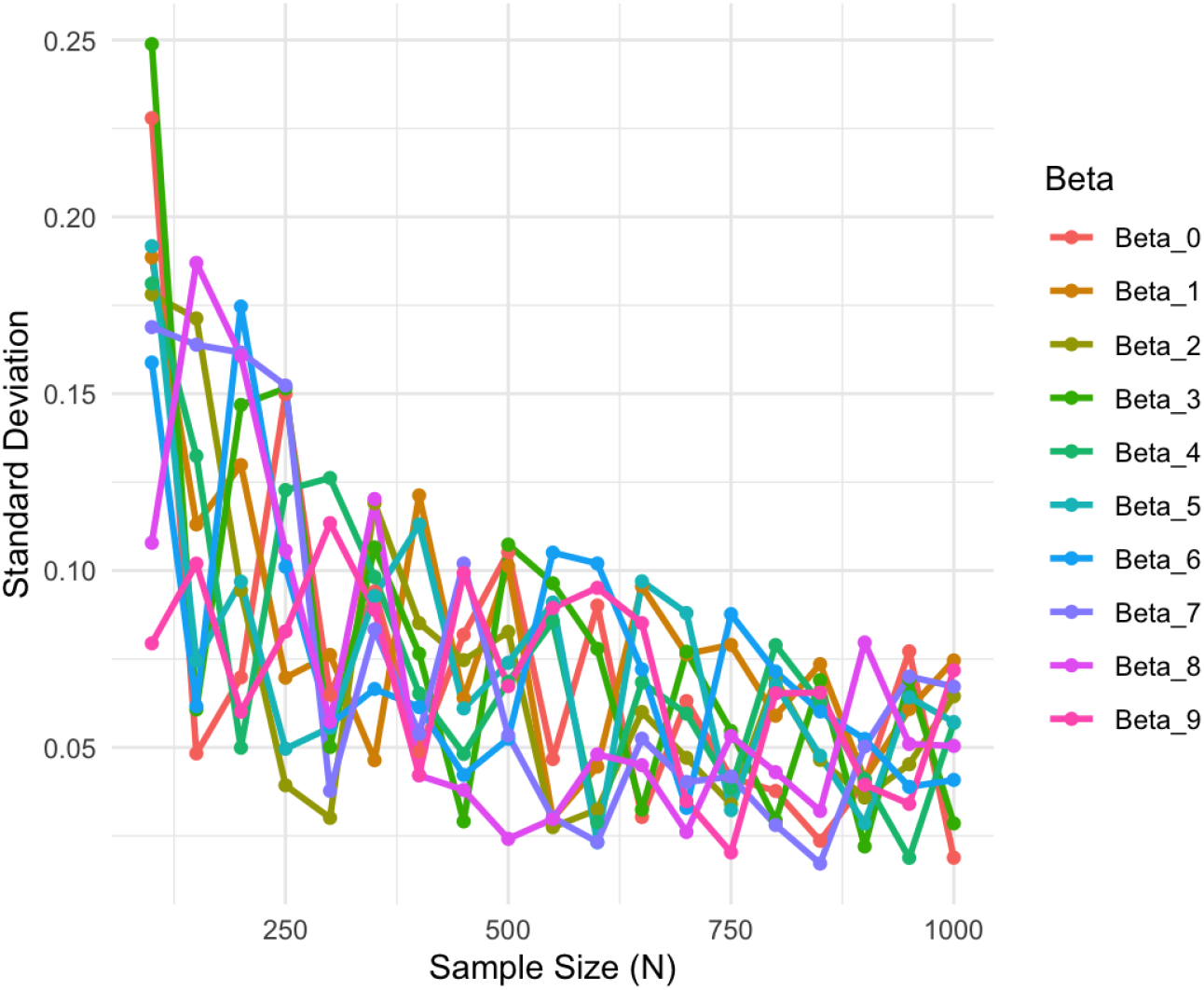
Variability of Estimated Parameters

Figure 10 illustrates the coefficient of variation (CV%) for the identical posterior estimates across the equivalent range of sample sizes. CV% quantifies the dispersion of parameter estimations by relating the standard deviation to the mean. As anticipated, the coefficient of variation (CV%) decreases with an increase in sample size. This indicates that the model exhibits greater accuracy and reduced uncertainty. However, with a reduced sample size, there is considerable volatility in CV%, with certain coefficients such as *β*_5_ and *β*_6_ exhibiting significant fluctuations. The anomalous results demonstrate how Bayesian inference can be influenced by inaccurate estimates in the absence of substantial data, underscoring the significance of an adequate sample size in hierarchical models.

**Figure 10.**
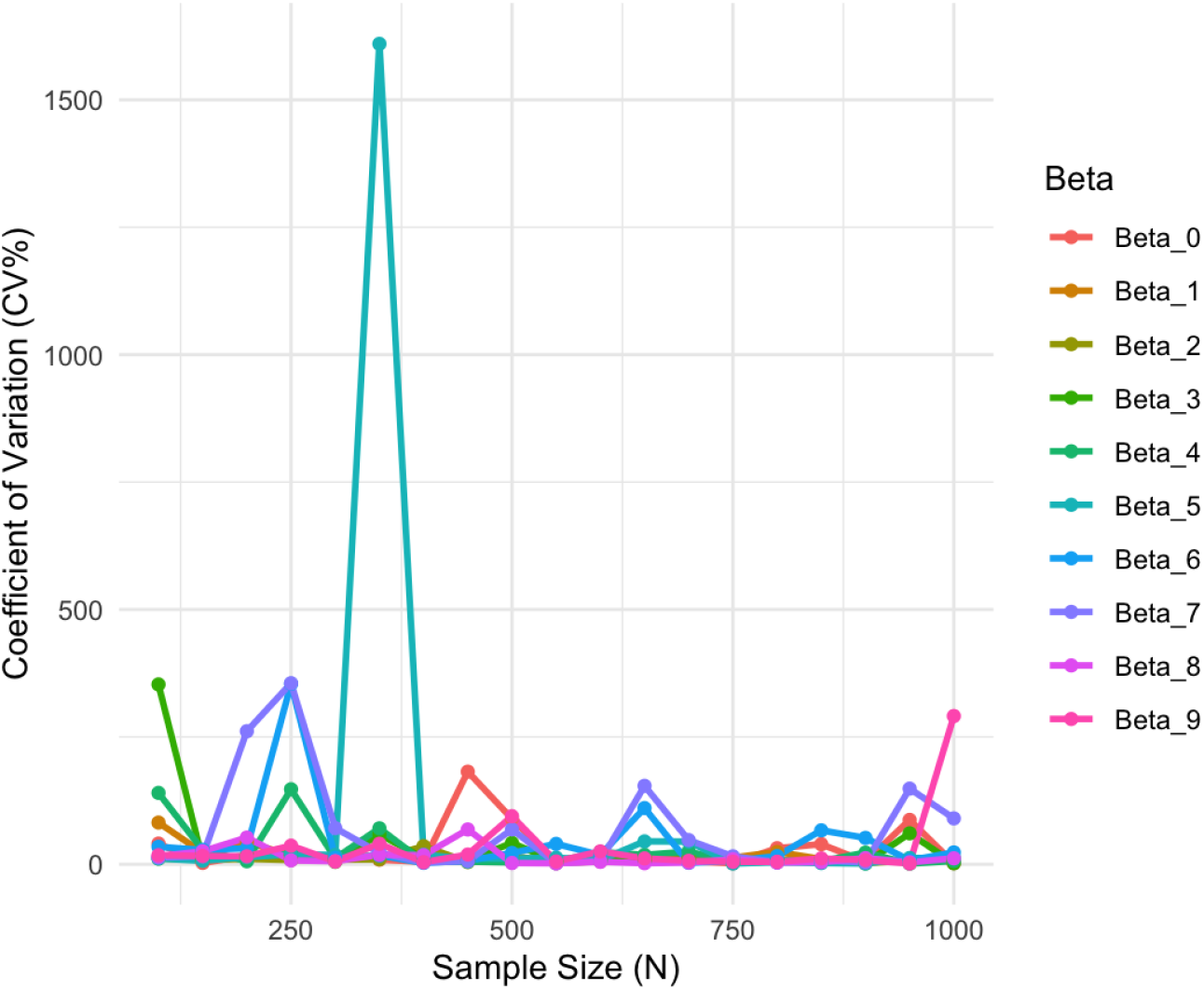
Variability of Estimated Parameters

The results indicate that the hierarchical Bayesian logistic regression model grows increasingly stable and reliable with the availability of additional data. The reduction in both the standard deviation and the coefficient of variation indicates that the estimates are converging towards the true parameter values, hence diminishing noise in the estimations. This empirical study demonstrates that Bayesian modeling is an effective method for analyzing health data, such as determining the prevalence of malnutrition among children, particularly when extensive data is accessible.

## 5 Discussion

This study demonstrates the significance of considering measurement error when analyzing pooled crosssectional survey data on health outcomes such as child malnutrition. The Bayesian adjusted model outperformed the uncorrected model across several performance parameters, including AUC, accuracy, and mean squared error (MSE). These modifications, however minor in many instances, provide a significant enhancement in the model’s capacity to discern distinctions and its general trustworthiness in predictions, particularly when misclassification is evident.

The uncorrected model consistently overestimates the malnourished population, particularly in later years such as 2022. This demonstrates that neglecting measurement error may result in inflated estimates of the public health cost, thereby leading to erroneous policy decisions and misallocation of resources. The revised model, conversely, provides prevalence estimates that are more conservative and likely more aligned with the actual underlying rates. This demonstrates how Bayesian approaches can enhance the reliability of health survey data.

The revised model has significantly narrower confidence bounds and posterior ranges, indicating enhanced precision of the parameters. This enhances the model’s robustness. Numerous elements in the model yielded significant posterior values, indicating their efficacy in predicting malnutrition risk. It is astonishing that the impacts of certain variables exhibit such significant disparities in magnitude and orientation. This illustrates the complexity of the interaction between socioeconomic and demographic factors and malnutrition patterns.

The findings are beneficial for healthcare systems in resource-limited countries such as Cameroon, where recall bias, reporting inaccuracies, and varying measurement methodologies may compromise data reliability. Bayesian hierarchical models effectively rectify errors by utilizing historical data on sensitivity and specificity. This is particularly advantageous for analyzing extensive survey data such as the Demographic and Health Surveys (DHS).

This work contributes to the expanding corpus of research focused on rectifying measurement mistakes in epidemiological investigations. This research expands upon the foundational studies conducted by Carroll et al. (2006), Gustafson (2004), and more recently, Wang et al. (2023). This study’s employment of simulation and synthetic data facilitates model validation; nevertheless, next research should investigate the efficacy of these methods on actual DHS datasets and in other domains where measurement quality is problematic.

However, certain limitations persist. The sensitivity and specificity values were predetermined rather than derived from the data, thus introducing bias if the assumptions are inaccurate. The brms interface facilitates the management of substantial computational expenses and convergence limitations in intricate models, while practical application may still provide challenges.

The posterior standard deviations (SD) and coefficients of variation (CV%) for ten regression coefficients (*β*_0_ to *β*_9_) in the hierarchical Bayesian logistic regression model with a sample size of *N* = 1000 are presented in Table 7. These measurements aid in determining the uncertainty and variability of each projected parameter relative to the others.

The intercept parameter (*β*_0_) has the most uncertainty, characterized by a standard deviation of 0.254 and a coefficient of variation of 26.2%. This indicates that the baseline log-odds exhibit greater variability than can be accounted for by the factors. This is common because intercept terms frequently account for variances that cannot be elucidated across clusters and contexts. Conversely, *β*_8_, *β*_7_, and *β*_6_ exhibited the lowest CV% values (below 17%), indicating that their estimations were more consistent and precise. This is probably due to their associated variables exhibiting stronger relationships with the outcome or superior measurement quality.

The majority of the other coefficients exhibit a substantial range of variability, with CV% values ranging from 17 to 22 percent. The results indicate that the hierarchical Bayesian model achieves an effective balance between flexibility and stability in estimates with a sample size of 1000. The low CV% values for all parameters indicate that the model effectively utilizes data from various years and regions through partial pooling, which is a significant advantage of hierarchical modeling Gelman et al. (2013).

Furthermore, employing CV% as a diagnostic instrument enhances the standard deviation by illustrating the degree of uncertainty relative to the posterior mean (McElreath, 2020). This is particularly significant when analyzing health data, since variations in scaling factors or prevalence rates can substantially alter outcomes, rendering straight standard deviation comparisons inaccurate. This study indicates that CV% is an effective method for identifying moderately unstable predictors.

These findings further demonstrate the utility of hierarchical Bayesian models in epidemiological research, particularly when analyzing complicated survey data that encompasses measurement error. Bayesian approaches provide a reliable approach to quantify uncertainty and exhibit greater robustness in data-scarce situations Wang et al. (2023). A detailed examination of parameter variations facilitates comprehension and ensures that the results drawn from the regression model are statistically sound and applicable for policy formulation.

This paper demonstrates the significance of Bayesian approaches in addressing misclassification and presents a scalable and replicable strategy applicable in various public health scenarios.

## 6 Conclusion

This study’s findings indicate that a hierarchical Bayesian logistic regression model effectively addresses measurement error in pooled cross-sectional health survey data. The proposed strategy significantly enhances the assessment of malnutrition prevalence and improves the model’s performance for area under the curve (AUC), accuracy, and mean squared error. This is accomplished by incorporating established criteria for sensitivity and specificity. The corrected model consistently outperformed the uncorrected one, particularly in generating more precise prevalence estimates and exhibiting superior predictive ability. These findings illustrate the peril of utilizing outdated survey data. This may result in skewed conclusions and inadequately informed public health measures. This Bayesian approach provides a robust and scalable means to enhance data reliability in resource-constrained environments. This strategy aids policymakers and researchers utilizing unreliable data sources in enhancing their comprehension. The methodology must be used to empirical datasets from various global regions, and subsequent research should explore methods to enhance the model for the direct estimation of misclassification parameters from the data.

## Data Availability

Data Availability The data underlying the results presented in this study are publicly available from the Demographic and Health Surveys (DHS) Program. The Cameroon DHS datasets for the years 2004, 2011, 2018, and 2022 can be accessed upon reasonable request from the DHS Program website at https://dhsprogram.com/data/. Researchers must create an account and submit a brief description of their project to obtain permission for data access. The DHS Program then grants access to de-identified datasets for academic and policy research purposes. All data used in this analysis were anonymized prior to access, and no individual identifying information is included. No new data were generated during this study.

## References

UNICEF. (2023). The State of the World’s Children 2023: For Every Child, Vaccination. UNICEF Reports. Retrieved from https://www.unicef.org/reports/state-worlds-children-2023

Victora, C. G., et al. (2021). Tracking development assistance for health and for COVID-19: a review of development assistance and funding mechanisms. The Lancet, 397 (10271), 1327–1337. 10.1016/S0140-6736(21)00073-0

Black, R. E., Victora, C. G., Walker, S. P., et al. (2013). Maternal and child undernutrition and overweight in low-income and middle-income countries. The Lancet, 382 (9890), 427–451. 10.1016/S0140-6736(13)60937-X

Smith, L. C., & Haddad, L. (2003). The Importance of Women’s Status for Child Nutrition in Developing Countries. IFPRI Research Report No. 131. https://www.ifpri.org/publication/importance-womens-status-child-nutrition

Rahman, M. M., et al. (2021). Determinants of stunting among children under 5 years in Bangladesh: evidence from a national survey. Public Health Nutrition, 24 (5), 1177–1190. 10.1017/S1368980020004204

Fuller, W. A. (2009). Measurement Error Models. John Wiley & Sons. https://www.wiley.com/en-us/Measurement+Error+Models-p-9780470371954

Gustafson, P. (2003). Measurement Error and Misclassification in Statistics and Epidemiology: Impacts and Bayesian Adjustments. CRC Press. 10.1201/9781420035484

Gelman, A., & Hill, J. (2006). Data Analysis Using Regression and Multilevel/Hierarchical Models. Cambridge University Press. 10.1017/CBO9780511790942

Carlin, B. P., & Louis, T. A. (2010). Bayesian Methods for Data Analysis (3rd ed.). Chapman and Hall/CRC. https://www.routledge.com/Bayesian-Methods-for-Data-Analysis/Carlin-Louis/p/book/9781439840955

Carroll, R. J., Ruppert, D., Stefanski, L. A., & Crainiceanu, C. M. (2006). Measurement Error in Nonlinear Models: A Modern Perspective (2nd ed.). Chapman and Hall/CRC. 10.1201/9781420010139

Everson, T. M., et al. (2020). Bayesian methods for correcting measurement error in epidemiological studies. Current Epidemiology Reports, 7, 60–71. 10.1007/s40471-020-00232-8

Ojomo, O., et al. (2022). Determinants of undernutrition among children under five in Nigeria: A multilevel analysis. BMC Public Health, 22, 1245. 10.1186/s12889-022-13644-7

Rahman, M. M., et al. (2020). Malnutrition and its associated factors among children under five in Bangladesh: A multilevel analysis. BMC Pediatrics, 20, 344. 10.1186/s12887-020-02269-5

Akombi, B. J., et al. (2017). Multilevel analysis of factors associated with wasting and under-weight among children under-five years in Nigeria. Nutrients, 9 (1), 44. 10.3390/nu9010044

Ngwira, A., et al. (2017). Child undernutrition in Malawi: A statistical analysis of dietary and socio-economic determinants. African Health Sciences, 17 (3), 684–695. 10.4314/ahs.v17i3.10

Cameroon National Institute of Statistics. (2023). Cameroon Multiple Indicator Cluster Survey (MICS). Retrieved from https://mics.unicef.org/

Lesaffre, E., & Lawson, A. B. (2020). Bayesian Biostatistics. John Wiley & Sons. https://www.wiley.com/en-us/Bayesian+Biostatistics-p-9780470018231

Silva, T., et al. (2022). Spatial modeling of child undernutrition in Sub-Saharan Africa: A Bayesian approach. Spatial and Spatio-temporal Epidemiology, 43, 100488. 10.1016/j.sste.2022.100488

Gelfand, A. E., et al. (2017). Bayesian Statistics: Principles, Models, and Applications. CRC Press. https://www.routledge.com/Bayesian-Statistics-Principles-Models-and-Applications/Gelfand-Schliep/p/book/9781498747114

Wakefield, J. (2013). Bayesian methods for hierarchical models and spatial data in epidemiology. In P. Damien et al. (Eds.), Bayesian Theory and Applications (pp. 393–433). Oxford University Press.10.1093/acprof:oso/9780199695607.003.0013

Banerjee, S., Carlin, B. P., & Gelfand, A. E. (2004). Hierarchical Modeling and Analysis for Spatial Data. Chapman and Hall/CRC. 10.1201/9781584887386

Li, X., et al. (2021). Bayesian spatial modeling of childhood stunting in low-resource settings: Evidence from Ethiopia. BMC Public Health, 21, 1358. 10.1186/s12889-021-11422-7

Gustafson, P. (2004). Measurement Error and Misclassification in Statistics and Epidemiology: Impacts and Bayesian Adjustments. Chapman and Hall/CRC. 10.1201/9780429134856

Wang, Z., Wang, Y., Li, X., & Wu, Y. (2023). Bayesian hierarchical modeling for misclassified binary outcomes in epidemiological studies. Statistical Methods in Medical Research, 32 (2), 175–192. 10.1177/09622802221136507

Gelman, A., Carlin, J. B., Stern, H. S., Dunson, D. B., Vehtari, A., & Rubin, D. B. (2013). Bayesian Data Analysis (3rd ed.). Chapman and Hall/CRC.

McElreath, R. (2020). Statistical Rethinking: A Bayesian Course with Examples in R and Stan (2nd ed.). CRC Press.

Spiegelhalter, D. J., Best, N. G., Carlin, B. P., & van der Linde, A. (2002). Bayesian measures of model complexity and fit. Journal of the Royal Statistical Society: Series B, 64 (4), 583–639. 10.1111/1467-9868.00353

Goldstein, H. (2011). Multilevel Statistical Models (4th ed.). Wiley.

